# Evaluative Stance Toward Artificial Intelligence in High-Quartile Medical Journals (2021– 2026): Large-Scale LLM-Assisted Computational Content Analysis

**DOI:** 10.64898/2026.07.23.26358815

**Authors:** Lixing Wang, Dan Poenaru

## Abstract

**Background:** Medical-AI publications do more than report technical performance; they also frame AI as beneficial, uncertain, or risky. How this evaluative stance has changed across the medical literature is not well characterized.

**Objective:** To characterize evaluative stance in published medical-AI discourse abstracts from January 2021 through April 2026 and examine variation over time, concern themes, failure mechanisms, specialties, first-author geography, and publication format.

**Methods:** We conducted an LLM-assisted computational content analysis of medical-AI abstracts from first- and second-quartile medical journals. Of 97,492 post-cutoff Q1/Q2 records entering the prefilter, 16,759 were retained as discourse or evaluative. Claude Sonnet 4.6 assigned 16,749 valid stance classifications using Alarm, Caution, Neutral, Cautious Optimism, and Advocacy. Annual analyses used 16,747 records dated 2021–2026. Critical stance was Alarm plus Caution and indexed evaluative scrutiny, rather than opposition or author psychology. Each LLM step was validated against blinded human coding by one author: prefilter Cohen κ=0.51, stance quadratic-weighted κ=0.79 (95% CI 0.72–0.84) for codable, in-scope records, specialty κ=0.75, and mechanism axes κ=0.84 (model type) and κ=0.57 (failure mode).

**Results:** Advocacy declined from 2.9% in 2021 to 0.6% in partial 2026, while Cautious Optimism remained the majority stance. Among 16,749 valid classifications, 30.8% were critical. Critical share increased from 25.4% to 32.6%, a 7.25-percentage-point increase based on unrounded estimates. Among critical records, patient safety remained the most prevalent concern. Hallucination/errors increased by 30.9 percentage points. Regulation declined by 22.0 percentage points and ethics/bias by 8.1 percentage points in prevalence share; these declines do not necessarily indicate lower publication counts. Within the hallucination/error theme, factual error was more common than fabrication. Fabrication estimates should be treated as an upper bound, because failure-mode agreement was moderate. Specialty patterns were heterogeneous. Critical rate was inversely associated with FDA cleared-device availability (Spearman ρ=−0.65, two-sided p=0.004), which does not measure adoption, deployment, maturity, or clinical use. First-author geography described publication metadata and discourse, not national attitudes or research quality. Reviews were the least critical and most favourable format. In exploratory forward-validation, 2 of 78 early Advocacy predictions were fully borne out, although the analysis was single-rater and retrieval-dependent.

**Conclusions:** Published medical-AI abstracts became modestly less promotional and more focused on specific errors and safety concerns. Unqualified promotion declined, but qualified favourable framing remained dominant, and the rise in critical stance was modest. Concern moved toward errors and patient safety, with factual error discussed more often than fabrication. These findings describe published discourse, not AI capability or whether the evaluations were correct.

## Introduction

Artificial intelligence (AI) has entered clinical medicine quickly, with machine-learning and deep-learning systems now reported across diagnosis, clinical decision support, risk prediction, medical education, and documentation (1–3). Its arrival has accelerated since late 2022, when general-purpose large language models (LLMs) were tested on clinical tasks such as licensing examinations and patient questions and were shown to encode substantial clinical knowledge (4, 5). Yet AI has entered the medical literature tentatively, with the same body of work that promotes AI also frequently worrying about it, often in the same abstract. Alongside optimism about efficiency and diagnostic performance runs a widening set of concerns: whether models are accurate enough for clinical use, whether the tendency of LLMs to produce confident but unreliable output (hallucination) can be contained (6), and whether patient safety can be assured. A parallel governance conversation asks how AI should be regulated as a medical technology (7, 8).

How the medical literature holds this tension is itself worth measuring. By “stance” we mean the evaluative position an article takes toward AI: whether it presents the technology mainly to promote it, or mainly to scrutinize it. Stance is a property of the published text, not of the authors’ private attitudes and not of how capable AI in fact is. It matters in its own right, because published discourse can shape how readily AI is promoted or criticized and which risks become visible, whatever the technology eventually proves able to do.

The medical-AI literature has itself been studied, but along dimensions that leave this stance uncharacterized. Bibliometric and scoping analyses have mapped the field’s growth, topics, and authorship rather than how it frames or criticizes AI (9, 10). Work closer to stance has measured sentiment, but outside the medical-journal literature: in public online discourse around the arrival of ChatGPT (11), and in the general valence, i.e., positive and negative word choice, of scientific abstracts (12), which connects to a broader concern with spin (reporting that overstates favourable conclusions beyond what the results support) and overinterpretation in biomedical writing (13). What no prior study has measured, to our knowledge, is whether the medical-AI journal literature itself promotes or scrutinizes AI, and how that balance has shifted across successive waves of the technology.

LLMs now make it feasible to close this gap, classifying scientific abstracts at scale at agreement approaching that of human annotators when the classifier is validated against human coding (14, 15). In a large-scale, LLM-assisted content analysis, we analysed AI-engaged abstracts from high-quartile (Q1/Q2) medical journals published between January 2021 and April 2026, assigning each abstract a stance with a classifier validated against blinded human coding. We read a critical stance as a broad index of evaluative scrutiny rather than editorial opposition, because the measure registers cautionary or negative findings even when they are reported dispassionately, and we examine it across both reflective writing about AI and system-testing writing that evaluates specific AI systems (i,.e. both discourse and evaluative genres). The current study contributes a validated stance classification, an explicit separation of fabrication from factual error in how papers describe AI failure, and an exploratory forward-validation of the field’s early optimistic (Advocacy) predictions. We characterize the stance these abstracts take toward AI and how it is distributed over time and across concern themes, failure mechanisms, specialty and domain categories, first-author geography, and publication format.

## Methodology

### Study Design and Corpus Construction

We conducted an LLM-assisted computational content analysis of medical-AI discourse in abstract-level records from Q1/Q2 medical journals (January 2021–April 2026). Bibliometric methods constructed the corpus but were not the primary study design.

We retrieved English-language PubMed/MEDLINE records monthly using AI-technology and medical-context terms. PubMed Publication Type metadata were collapsed into five categories: Research Article, Editorial, Review, Commentary, and Letter. Query, mapping rules, and code are in the code repository (Supplementary Note 1). Retrieval yielded 130,181 records and 116,117 unique PubMed identifiers after deduplication. Excluding 4,106 conference proceedings and preprint records left 112,011 for annual SCImago Journal Rank matching by normalized journal title. SJR best quartile determined inclusion; journals qualified if Q1/Q2 in any subject category. Of these records, 98,626 were Q1/Q2 (88.0%), 7,922 were unmatched, and Q3 was retained as a comparison arm using the same v1 rubric (Figure S5, Table S12). Applying the April 2026 cover-date cutoff reduced Q1/Q2 records from 98,626 to 97,492. The 2026 stratum covered January–April and was included in descriptive six-year trend summaries, with its partial coverage noted throughout.

### AI-Engagement Prefilter

Claude Haiku 4.5 (claude-haiku-4-5-20251001, Batch API; temperature not explicitly set, provider default T=1.0) classified titles and abstracts as discourse, evaluative, or application. Discourse records discussed AI; evaluative records tested a system and assessed accuracy, safety, or readiness; application records used AI only as a method. We retained discourse and evaluative records and excluded application records. Parsing and failure rules are in the code repository (Supplementary Note 1). We retained 16,759 records: 8,899 discourse and 7,860 evaluative.

Each LLM classification step was validated against human coding. One author with AI research experience coded every validation sample, blind to the model’s labels. Agreement is reported as Cohen κ, quadratic-weighted for the ordered stance scale and unweighted for the nominal axes, with percentile bootstrap CIs (n=1000, seed=42). In a 200-record prefilter pilot, raw agreement was 85.5% and Cohen κ was 0.51 (95% CI 0.35–0.68). The annual keep rate, defined as the proportion of eligible Q1/Q2 records retained as discourse/evaluative records, was used to audit temporal filtering (Table S15). Specialty-specific keep rates could not be estimated because excluded application records lacked specialty labels–specialty comparisons were therefore treated as exploratory.

### Stance Classification and Validation

Claude Sonnet 4.6 (claude-sonnet-4-6, Batch API; temperature not explicitly set, provider default T=1.0) classified each title and full abstract with canonical v1 prompt as Alarm, Caution, Neutral, Cautious Optimism, or Advocacy. Critical stance was Alarm plus Caution; Favourable stance was Cautious Optimism plus Advocacy. Critical stance indexed evaluative scrutiny in the abstract, including dispassionate underperformance reports, not an explicit editorial opposition or author attitudes. Classification emphasized the authors’ discussion and conclusion over positive background framing (code repository; Supplementary Note 1).

There were 16,749 valid classifications; 10 unparseable responses were dropped. Temporal and stratified analyses used 16,747 records dated 2021–2026 after excluding two dated 2020. Validation used 300 records stratified by assigned stance, with Alarm and Advocacy oversampled. Quadratic-weighted Cohen κ was 0.79 (95% CI 0.72–0.84) for codable, in-scope records (n=225), 0.75 (0.69–0.81) across all 300, and 0.80 after additionally excluding survey or meta-discourse records (n=218).

A stricter conclusion-anchored v2 prompt tested sensitivity; v1 remained canonical throughout (Table S2). Run-to-run stability at T=1.0 used two re-inferences of the 300 validation records. Agreement was 95.3%, with quadratic-weighted κ=0.97; all 14 changes were adjacent, with no polar flips (Table S16).

### Theme and Mechanism Classification

Among 5,161 critical records, Claude Sonnet 4.6 (claude-sonnet-4-6, Batch API; temperature not explicitly set, provider default T=1.0) assigned one to three themes from the title and first 400 abstract characters: safety_clinical, hallucination, regulation, ethics_bias, education, data_privacy, cognitive, existential, replacement, and other. Verbatim definitions are in the code repository (Supplementary Note 1).

Because hallucination included inaccurate and fabricated outputs, Claude Sonnet 4.6 (claude-sonnet-4-6, Batch API; temperature explicitly set to 0.0) classified two axes. Failure mode was confabulation, misclassification, both, or none_or_unclear; model type was generative, discriminative, both, or unclear. Confabulation is reported as fabrication and misclassification as factual error. Analyses covered the hallucination-gated subset and full abstract-present critical corpus. Of 5,161 records, 5,159 were parsed; 2 were dropped. Excluding title-only records left 4,747.

Validation used 120 records stratified by year and assigned failure mode. Agreement used 108 records with abstracts after excluding 12 title-only records. For model type, Cohen κ=0.84, Gwet AC1=0.89, raw agreement=90.7%, and κ/κ_max=0.86. For failure mode, κ=0.57, AC1=0.60, raw agreement=69.4%, κ_max=0.76, and κ/κ_max=0.76. A nonblind disagreement review found more model-assigned fabrication labels, so fabrication estimates were treated as an upper bound (code repository; Supplementary Note 1).

### Specialty, Geography, and FDA Variables

Claude Haiku 4.5 (claude-haiku-4-5-20251001, Batch API) assigned one of 18 specialty or domain categories from titles and abstracts, based on clinical domain (Multimedia Appendix 1.7). Validation used 120 stratified records with an “unclear from abstract” option. Agreement was Cohen κ=0.75, Gwet AC1=0.75, raw agreement=76.7%, and κ/κ_max=0.89. Primary analyses excluded categories with fewer than 30 records. Supplementary mechanism analyses retained all categories, flagged those with fewer than 20, and suppressed their CIs.

First author and affiliation came from PubMed XML document order, without corresponding-author logic. Country was resolved by rule-based substring lookup and grouped into eight regions; 90.3% of records resolved. Maps required ≥50 records per country, with an n≥20 sensitivity map (Figure S4). Geography indexes first-authored publication metadata, not national attitudes or research quality. Mapping and ambiguity rules are in the code repository (Supplementary Note 1). Affiliation was also classified and analysed as industry, academic, mixed, or unknown (Table S13).

Cleared-device availability used the U.S. FDA list of AI/ML-enabled medical devices [accessed on June 2, 2026, https://www.fda.gov/medical-devices/software-medical-device-samd/artificial-intelligence-enabled-medical-devices], comprising 1,430 devices cleared between 1995 and 2025. Lead review panels were mapped to the 18 specialties. Specialties without a corresponding panel received structural zeros, meaning no mapped FDA panel, not an absence of clinically available AI in the field. The mapping is in the code repository (Supplementary Note 1). Generative-share was the proportion of each specialty’s critical papers classified as generative or both.

### Forward-Validation of Early Predictions

This exploratory analysis began with all 92 Advocacy records obtained from 2021–2023 carrying a forward-looking predictive-claim flag. Claude Sonnet 4.6 (claude-sonnet-4-6, temperature=0) extracted the most falsifiable prediction and assigned Tier 1 (specific superiority or replacement), Tier 2 (a concrete capability entering clinical use), or Tier 3 (diffuse transformation with no mechanism or horizon). One author reviewed source abstracts blinded to later evidence. Removing 18 false positives gave post-hoc flag precision 0.80 and a frozen set of 74 papers with 78 claims (four papers contained an independent second claim), distributed Tier 1 = 7, Tier 2 = 36, Tier 3 = 35.

Claims were matched to 58,203 abstracts published in 2024–2026 from the broad Q1/Q2 corpus, including all application records previously excluded for other analysis. MedCPT bi-encoder dot-product similarity returned the top 20 abstracts per claim. Claude Sonnet 4.6 (temperature=0) summarized retrieved evidence but assigned no verdict. One author assigned borne out, partially borne out, not borne out, or too early/unfalsifiable and cited at least one evidence PMID per verdict. Not borne out required positive contrary evidence. Corpus silence or retrieval below the prespecified similarity threshold was classified as too early/unfalsifiable. The tier taxonomy and adjudication rubric were frozen before verdicts. The frozen protocol is in the code repository (Supplementary Note 1); the Tier-by-verdict matrix is in Table S11. It was Advocacy-only, single-rater, retrieval-dependent, and limited by the April 2026 cutoff.

### Statistical Analysis and Reproducibility

Proportions and rates used 95% percentile bootstrap CIs from 1000 row-level resamples with replacement (seed 42). Six annual estimates from 2021–2026 were summarized by Spearman correlation, with year as a descriptive monotonic-trend measure. Publication-type distributions were compared using a χ² test of independence and Cramér’s V (Table S8).

Across specialties, critical rate was compared with FDA cleared-device availability using Spearman correlation. The two-sided p value was primary; one-sided and permutation variants were sensitivity analyses. We repeated the analysis after removing structural-zero categories. Zero-order Spearman correlation compared critical rate with generative-share. Because generative-share was calculated among critical papers, adjustment for it was treated as a sensitivity analysis (Table S5).

Temporal critical share was repeated within discourse and evaluative records (n=16,747), using the same bootstrap CIs and descriptive Spearman correlations. Annual discourse:evaluative ratios assessed composition (Figure S6, Table S14). Q3 and prefilter sensitivity analyses are in Figure S5 and Tables S12 and S15.

Analyses were descriptive and hypothesis-generating. P values were not used for confirmatory decisions or adjusted for multiplicity; interpretation emphasized effect sizes and CIs. Analyses used Python 3.12, with package versions pinned in the repository. Prompts, code, PubMed identifiers, and committed outputs are available [https://github.com/Lixing-Kingsley-Wang/ai-narratives-2026]. Hosted-model drift makes committed outputs the basis for exact reproduction. The study involved no human participants or identifiable patient data; institutional ethics review was not required.

## Results

### Corpus and Overall Stance

Of 97,492 Q1/Q2 records after the April 2026 cutoff, the prefilter retained 16,759 AI-engaged records: 8,899 discourse and 7,860 evaluative records. Stance classification yielded 16,749 valid labels after 10 unparseable responses were dropped. Temporal and stratified analyses used 16,747 records dated 2021–2026; two records dated 2020 were excluded. Overall, 30.8% were critical, comprising 916 Alarm and 4,245 Caution records, and 69.2% were non-critical. Cautious Optimism formed most of the non-critical majority, while Advocacy was rare (n=235; Figure 1A).

**Figure 1.**
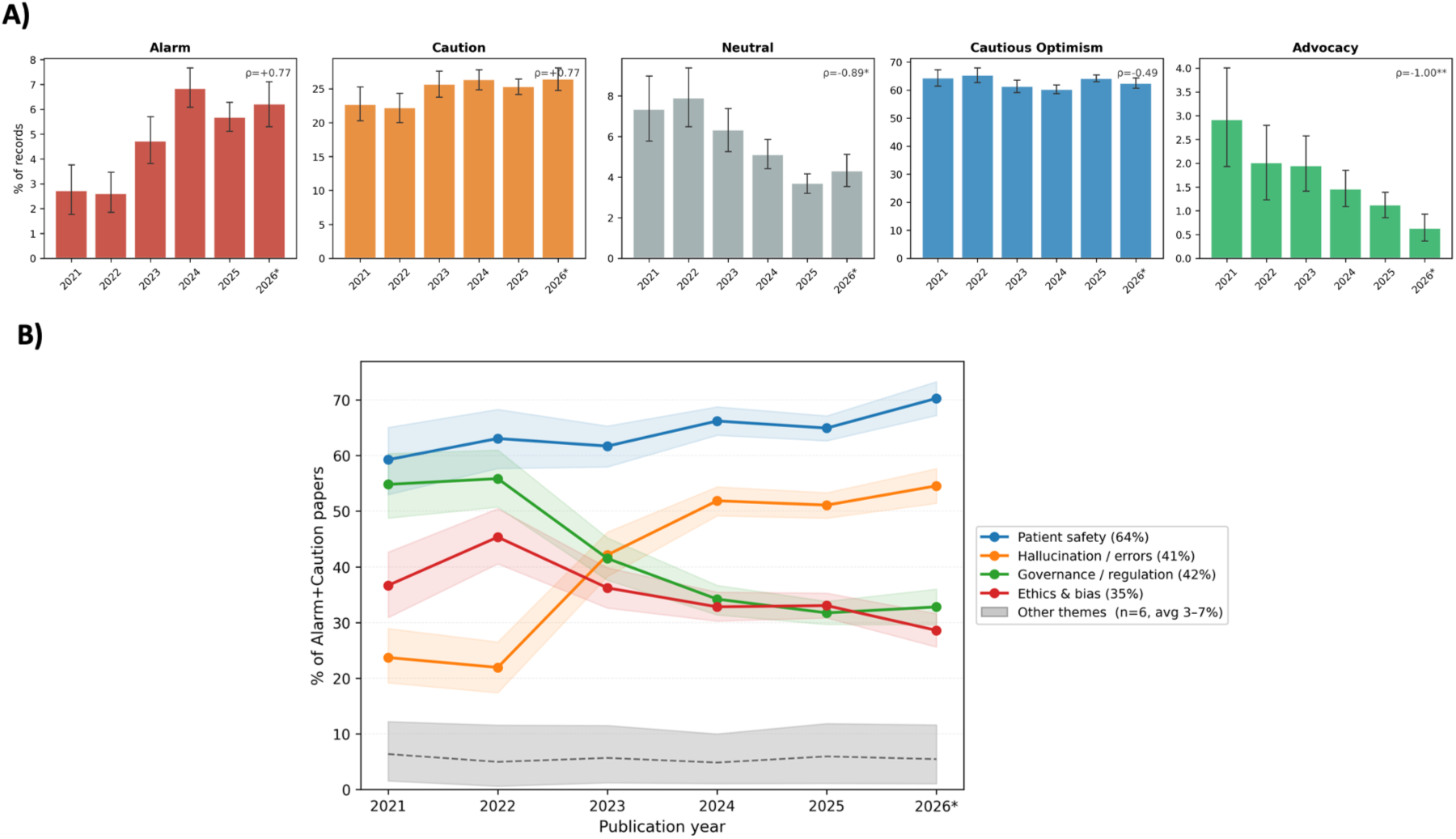
Temporal distribution of AI stance and concern themes in high-quartile medical journals, 2021–2026. (A) Annual distribution of Alarm, Caution, Neutral, Cautious Optimism, and Advocacy among 16,747 records with valid 2021–2026 publication years, from 16,749 valid-stance records after excluding two dated 2020. Critical stance was Alarm or Caution; favourable stance was Cautious Optimism or Advocacy. Bars show annual estimates with 95% percentile bootstrap CIs (1000 resamples, seed=42). Spearman ρ summarizes association with publication year. (B) Annual prevalence of four leading themes among critical records (n=5161). Themes were multilabel, with one to three per paper, so prevalence does not sum to 100%. Lines show annual estimates; shaded bands show 95% percentile bootstrap CIs. The six lower-prevalence themes are grouped as “other themes.” Percentages in the panel legend are mean annual prevalences. The 2026 stratum is partial (January–April). *p<.05; **p<.01 by exact two-sided permutation.

### Temporal Change and Concern Themes

Critical share rose from 25.4% [22.9–28.1] in 2021 to 32.6% [30.9–34.4] in the partial 2026 stratum, a net increase of 7.25 percentage points (Figure 1A). The direction was preserved under the stricter v2 rubric (Table S2). The rise was gradual, with no peak in 2023 or subsequent reversion. Alarm rose from 2.7% in 2021 to 6.8% in 2024, and was 6.2% in partial 2026; Caution rose from 22.7% to 26.4%. Cautious Optimism remained the majority every year. The January–April 2026 stratum was partial, and is therefore reported descriptively. Advocacy fell from 2.9% in 2021 to 0.6% in 2026 (Spearman ρ=−1.00). Of the 17 Advocacy records in 2026, 5 were explicitly about drug discovery. The remaining Advocacy was concentrated in methodology and drug-discovery papers; clinical-specialty Advocacy had all but disappeared. The record-level breakdown is in Table S3.

The temporal rise was present within both prefilter genres among the 16,747 year-dated records. Critical share in discourse papers rose from 30.2% to 37.8% (Spearman ρ=+0.94), while the evaluative-paper share rose from 14.6% to 28.1% (ρ=+0.77). The latter trend was descriptive, and not conventionally significant across six annual points. Meanwhile, the discourse:evaluative ratio fell from 2.24 in 2021 to 0.88 in 2026. The overall rise was therefore not only due to changing genre composition (Figure S6, Table S14). The annual keep rate rose monotonically over the period (Spearman ρ=+1.00; Table S15), making differential filtering an unlikely source of the trend.

Concern themes could overlap; each of the 5,161 critical records was assigned one to three themes. Annual prevalence used the number of critical papers in each year as the denominator. Four themes carried the main temporal changes in Figure 1B; the other six are shown in Figure S1. Patient safety remained the most prevalent theme and rose by +11.0 percentage points (Figure 1B). The hallucination/errors theme rose by 30.9 percentage points and became the second most prevalent concern. Regulation fell by 22.0 percentage points, while ethics/bias fell by 8.1 percentage points, from 36.7% in 2021 to 28.6% in 2026. These were declines in prevalence share. The absolute number of regulation-themed papers remained roughly steady as the critical corpus expanded, so the lower prevalence reflected dilution rather than a fall in volume. Full theme trends and per-theme Spearman results are in Figure S1 and Table S1; counts and prevalence shares are in Figure S2.

### Failure Mechanisms

Mechanism classification parsed 5,159 of the 5,161 critical records; 2 unparseable records were dropped. Excluding title-only records left 4,747 abstract-present critical records. The failure-mode axis distinguished confabulation, reported as fabrication, from misclassification, reported as factual error. It also included both and none_or_unclear. The model-type axis distinguished generative, discriminative, both, and unclear systems.

Within the hallucination-gated subset of 2,226 abstract-present records, factual error was the majority mechanism in every year, accounting for 56% to 90% of gated papers (Figure 2A). Fabrication-involved papers, defined as fabrication alone or with factual error, rose from 0% in 2021 to roughly 21–23% from 2023 onward. The previous “hallucination” thematic gate captured about nine in ten fabrication-related papers, but only about one in five of the gated papers concerned fabrication. Its annual composition therefore remained dominated by factual error, even after fabrication-related papers increased. Therefore, previous hallucination label did not denote fabrication alone.

**Figure 2.**
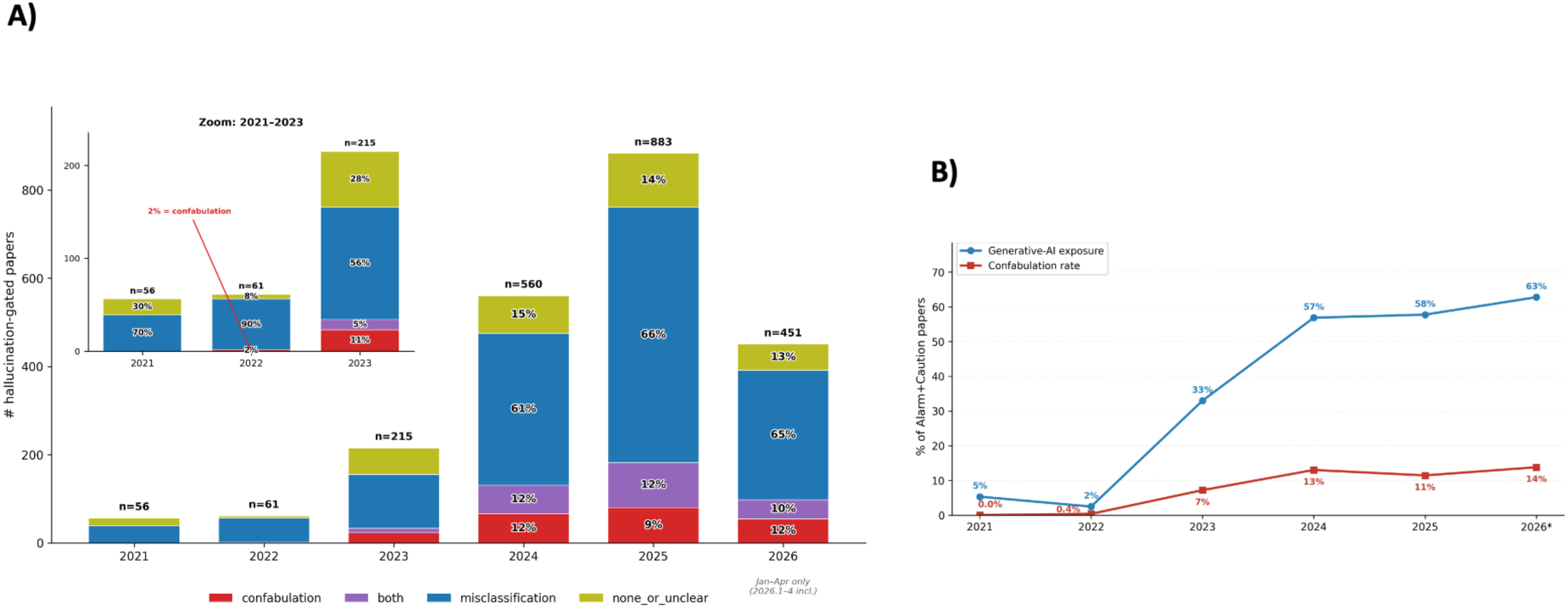
Failure-mode and model-type composition of hallucination-flagged critical records. (A) Annual failure-mode composition among hallucination-gated, abstract-present critical records (n=2226). Critical stance was Alarm or Caution. Bar height shows the yearly record count; segments show within-year proportions classified as confabulation, misclassification, both, or none/unclear. Confabulation is reported as fabrication, and misclassification as factual error. Each bar sums to 100%. (B) Annual generative-model exposure and confabulation rate in the full abstract-present critical corpus (n=4747). Generative-model exposure is the proportion classified as generative or both; confabulation rate is the proportion classified as confabulation or both. The hallucination gate includes factual error and fabrication. The 2026 stratum is partial (January–April).

Across all 4,747 abstract-present critical records, the generative-model share rose from 5.3% in 2021 to 62.8% in partial 2026 (Spearman ρ=+0.94; Figure 2B). Fabrication share also rose monotonically (ρ=+0.94) but plateaued in the low teens, reaching 13.8% in 2026. Among generative-model papers, fabrication remained near 20–23% across the period (Table S4). In total, 1,858 records (39.1%) concerned generative models without a fabrication concern; most instead assessed accuracy, bias, or judgment. Only one paper raised fabrication in a non-generative discriminative system. Model type and failure mode therefore represented separate dimensions of the critical records. Because failure-mode agreement was moderate (κ=0.57) and the classifier assigned fabrication more readily than the human coder, fabrication estimates were treated as an upper bound.

### Specialty and Cleared-Device Availability

Specialty analyses included 16,734 records with valid assignments across 18 categories, each with at least 30 records; 13 records with failed specialty classification were excluded. Critical rate varied roughly threefold (Figure 3A). It was highest in Mental Health/Psychiatry at 51.3% (46.8–55.5), Medical Informatics/Digital Health at 44.9% (43.5–46.4), and Nursing at 43.0% (37.0–48.8). It was lowest in Cardiology at 16.8% (14.1–19.8), Ophthalmology at 18.4% (15.7–21.2), and Oncology at 19.7% (17.8–21.8).

**Figure 3.**
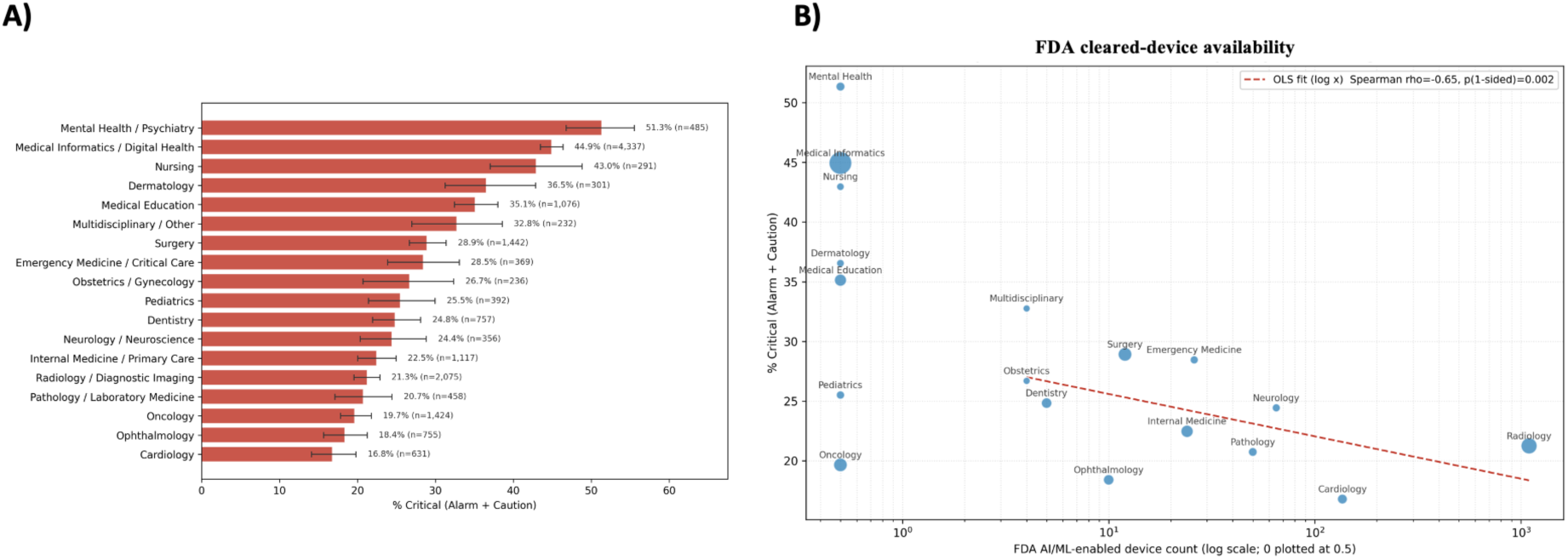
Critical stance by specialty/domain category and FDA cleared-device availability. (A) Critical rate among 16,734 records with valid specialty assignments; 13 records with failed classification were excluded. Critical stance was Alarm or Caution. Bars show category estimates with 95% percentile bootstrap CIs. (B) Critical rate versus FDA-cleared AI/ML device count across the same 18 categories. Each point represents one category, with size proportional to its record count. The x-axis is logarithmic; structural-zero categories with no mapped FDA device panel are plotted at 0.5. The dashed line shows the displayed OLS fit. FDA counts index US cleared-device availability, not adoption, deployment, maturity, or clinical use. The association is cross-sectional and does not establish a mechanism.

Critical rate was inversely associated with FDA-cleared AI/ML device count, used as a US-based measure of cleared-device availability (Spearman ρ=−0.65, two-sided p=0.004; Figure 3B). Radiology, Cardiology, and Pathology had among the largest mapped device counts and lower critical rates. Excluding the seven structural-zero categories produced a directionally similar estimate (ρ=−0.58, n=11), but it was no longer conventionally significant (two-sided p=0.062; Table S5). The one-sided and permutation analyses are also reported in Table S5. Critical rate was uncorrelated with generative-share (ρ=−0.01; Figure S3), with the partial-correlation analysis reported in Table S5. These cross-sectional associations did not establish a mechanism.

### First-Author Geography

Critical rate varied by first-author country (Figure 4). Papers with a United States first author were critical in 34.5% [33.0-36.0] of cases (n=4,222), compared with 16.2% [14.6-17.8] for China (n=1,904). Rates were at or above 40% in several northern and western European countries and Australia, including the Netherlands at 44.4%, Denmark at approximately 42.6%, Australia at approximately 42.5%, Ireland at 41.1%, and the United Kingdom at approximately 40.6%. Rates were near or below 20% in several East and South Asian countries and parts of the Middle East, including Japan at 18.3%, Iran at 18.0%, South Korea at 20.5, and India at 20.6%.

**Figure 4.**
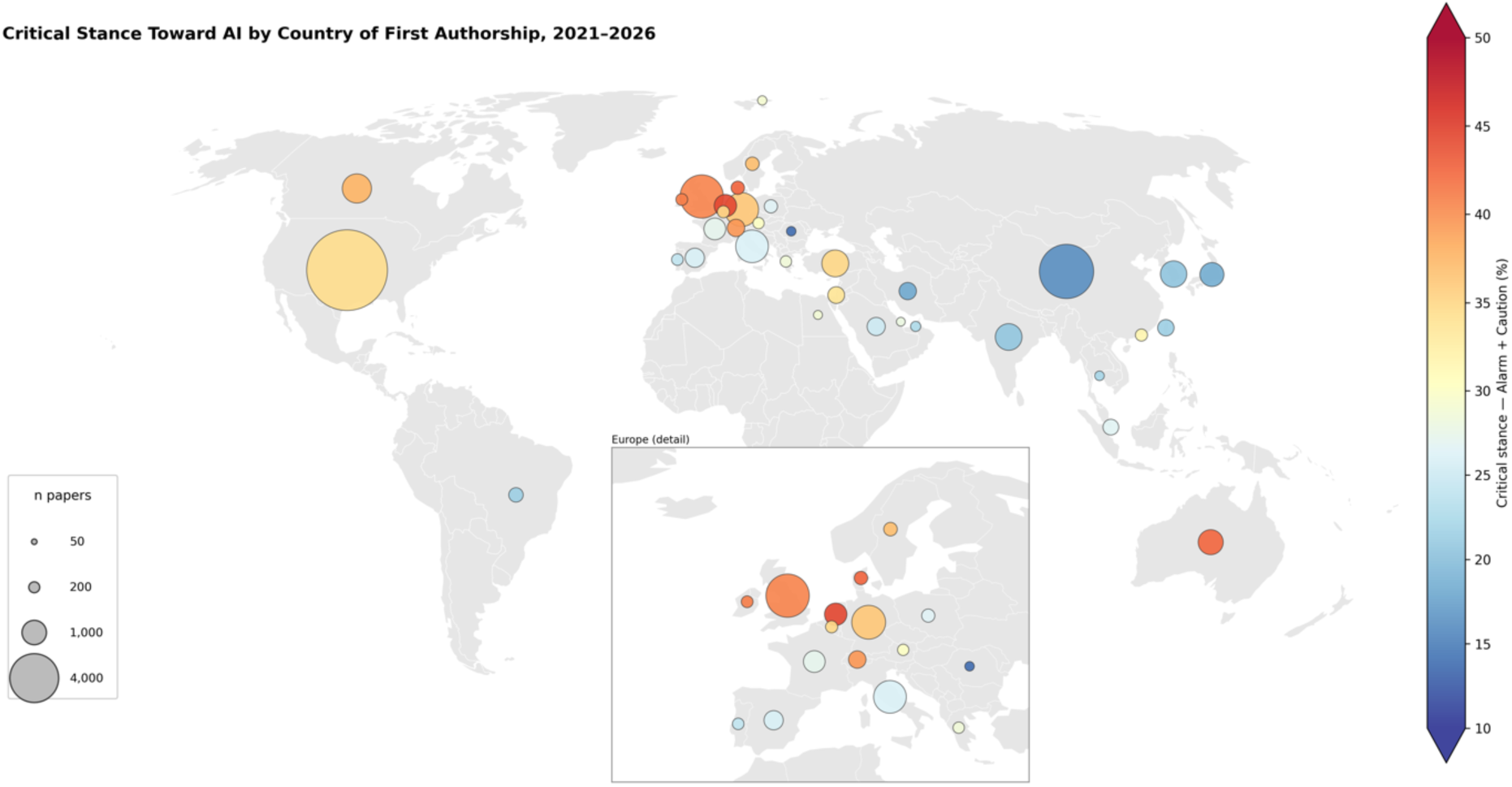
Critical stance toward AI by country of first authorship, 2021–2026. The map shows 36 countries with at least 50 first-author records. Bubble area is proportional to the number of records; fill colour shows the critical rate, defined as Alarm or Caution. Country was assigned from the first-author affiliation and indexes publication discourse, not national attitudes, research quality, or AI capability. The map uses a Robinson projection with a European inset. Additional country and regional analyses are in Multimedia Appendix 2. The 2026 stratum is partial (January–April).

North America and Western Europe both had first-author critical rates of 35.1% [33.9-36.4](n=4961) and [33.6-36.5](n=4296), respectively. East Asia had the lowest regional rate, 18.5% [17.2–19.9] (n=3,260), with a comparatively narrow confidence interval because of its large sample size. Reweighting East Asia’s specialty mix to match North America and Western Europe raised its rate from 18.5% to 19.9%, compared with 35.1% for North America and Western Europe, closing only 8% of the gap. Within East Asia, rates were 31.6% in Hong Kong, 26.8% in Singapore, and 16.2% in China. Per-country and per-region estimates are reported with Figure 4. The more permissive country map is in Figure S4, and East Asia comparisons are in Tables S6 and S7. Naturally, these results only index first-authored publication discourse, not national attitudes, research quality, or AI capability.

## Publication Type

Publication-type analyses used 16,747 records: Research Articles (n=10,315), Reviews (n=5,444), Letters (n=480), Editorials (n=404), and Commentaries (n=104). Critical rates were 53.9% for Commentaries, 42.1% for Letters, 38.4% for Editorials, 32.9% for Research Articles, and 24.9% for Reviews (Figure 5).

**Figure 5.**
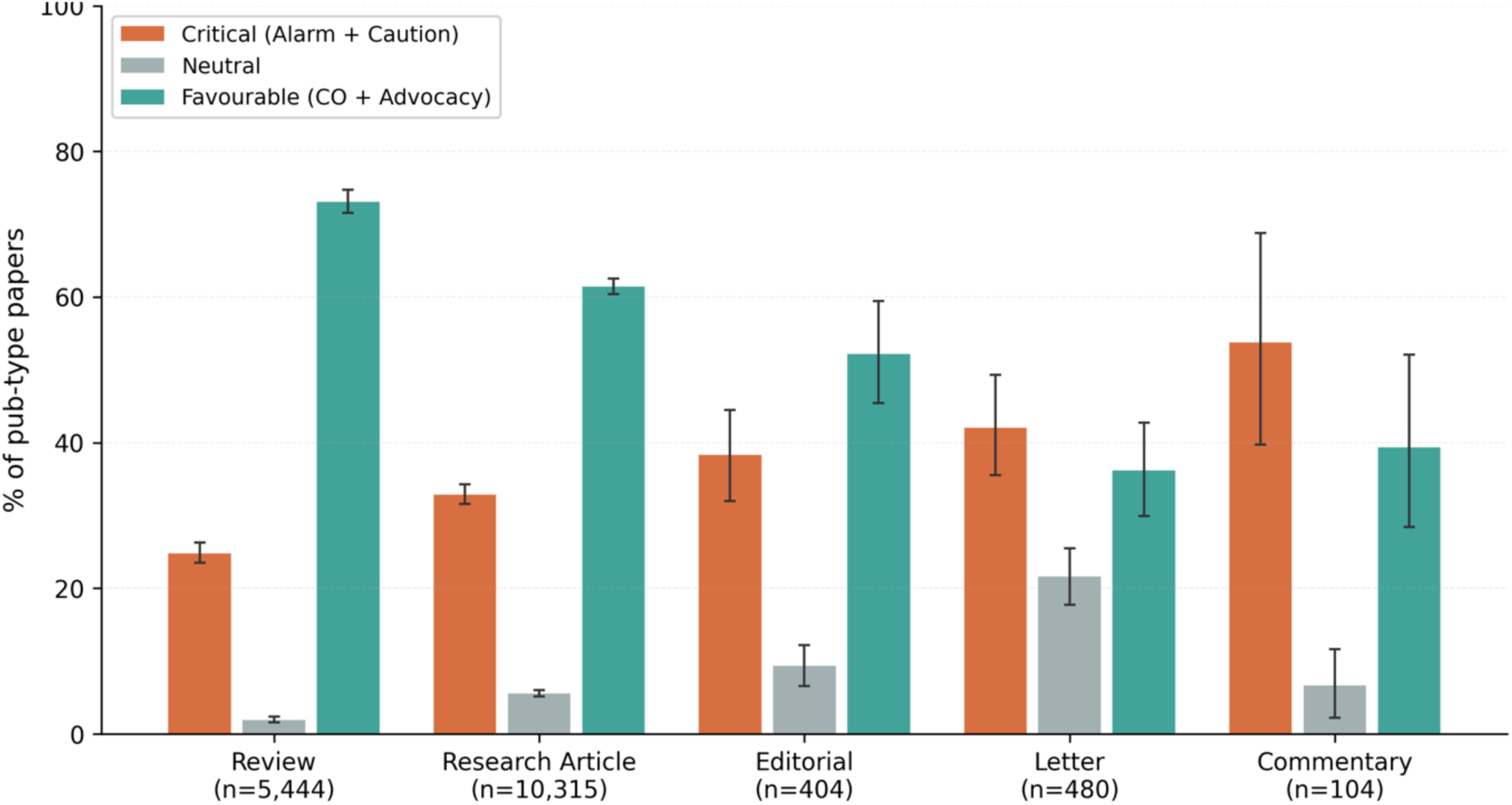
Stance distribution by publication type. Among 16,747 records dated 2021– 2026, bars show the proportion within each publication type classified as critical, neutral, or favourable. Critical stance was Alarm or Caution; favourable stance was Cautious Optimism or Advocacy. Publication-type sample sizes are shown below the x-axis labels. Error bars show 95% CIs. The full publication-type-by-stance table and related analyses are in Multimedia Appendix 2 (Tables S8–S10).

Pooling Commentaries, Letters, and Editorials, opinion formats were more critical than Research Articles (41.8% vs 32.9%). Their five-category stance distributions differed (N=11,303; χ²₄=430.5, p<0.001; Cramér’s V=0.195, a small effect; Table S8). Letters also had a neutral share of approximately 22%, compared with approximately 2–9% in most other formats.

Reviews were the most favourable format at 73.1%, followed by Research Articles at 61.5%, Editorials at 52.2%, Commentaries at 39.4%, and Letters at 36.3%. Reviews were therefore both least critical and most favourable. Systematic reviews were more critical than narrative reviews (29.3%, n=980 vs 23.9%, n=4,464; Table S9), while the larger narrative subtype accounted for most review records. The Commentary estimate was based on n=104 and had a correspondingly wide interval.

## Exploratory Forward-Validation

The exploratory forward-validation began with 92 Advocacy papers from 2021–2023 that carried a forward-looking predictive-claim flag. Blind human review removed 18 false positives, giving a post-hoc flag precision of 0.80. The frozen set contained 74 papers and 78 claims: Tier 1, n=7; Tier 2, n=36; and Tier 3, n=35 (Figure 6). A single rater compared each claim with the 2024–2026 published record. “Not borne out” required positive contrary evidence, while corpus silence was classified as too early/unfalsifiable.

**Figure 6.**
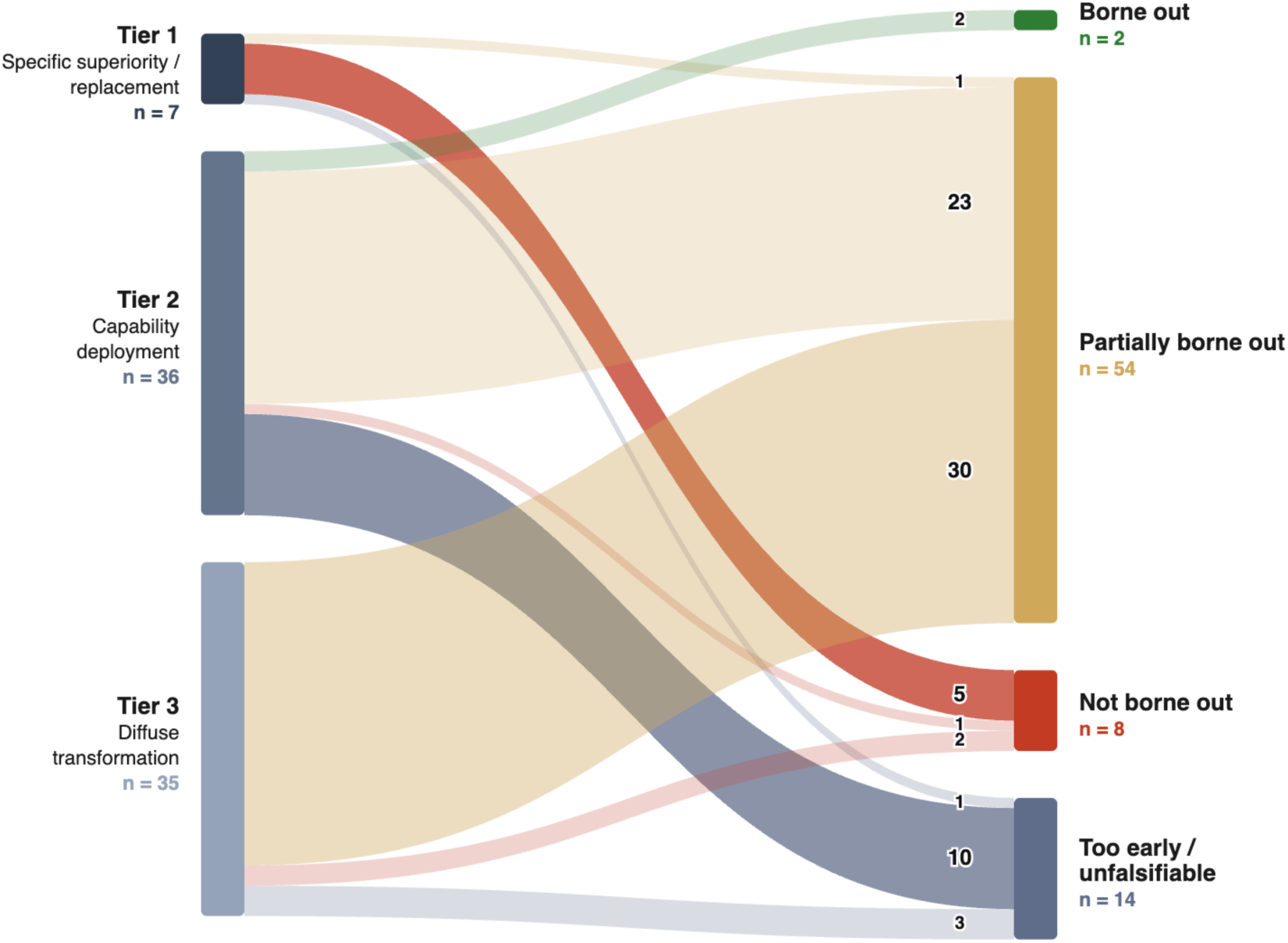
Exploratory forward-validation of early Advocacy predictions. The flow diagram shows 78 predictions from 74 Advocacy papers published in 2021–2023, grouped by falsifiability tier and adjudicated against the 2024–2026 evidence corpus. Node height and ribbon width represent claim counts. A single rater assigned each claim as borne out, partially borne out, not borne out, or too early/unfalsifiable. “Not borne out” required positive contrary evidence; corpus silence was classified as too early/unfalsifiable. The full Tier × verdict matrix is in Multimedia Appendix 2, Table S11. This analysis was exploratory, Advocacy-only, single-rater, retrieval-dependent, and limited by the April 2026 evidence cutoff. It does not adjudicate AI capability or whether critical stances were correct.

Of the 78 claims, two, both Tier 2, were borne out; 54 (69.2%) were partially borne out; eight were not borne out; and 14 were too early/unfalsifiable. Partially borne-out claims had moved in the predicted direction but were narrower, weaker, or slower than stated. Among Tier 1 specific-superiority or replacement claims, five out of seven were not borne out, the largest single not-borne-out flow, and none were borne out. Tier 2 capability-deployment claims were mainly partially borne out (23 of 36) and included both borne-out claims. Tier 3 diffuse-transformation claims were mainly partially borne out (30 of 35; Table S11). This secondary Advocacy-only, single-rater, retrieval-dependent analysis was limited by the April 2026 cutoff and does not generalize to all medical-AI forecasts or indicate that critical stances were correct.

## Discussion

### Principal Findings

From January 2021 to April 2026, evaluative stance in high-quartile medical-journal abstracts shifted in register and emphasis. Unqualified Advocacy declined sharply, but Cautious Optimism remained the majority stance and the overall rise in critical stance was modest. The increase appeared within both discourse and evaluative papers, so it was not solely caused by changing genre composition. Within critical papers, concern shifted toward errors and patient safety, while regulation and ethics/bias declined in prevalence share - though these declines do not imply fewer publications in absolute terms. Within the hallucination theme, factual error was more common than fabrication. Generative-model papers became much more common, but fabrication remained a minority concern, and its estimated prevalence should be treated as an upper bound. Scrutiny also varied across specialties, first-author countries, and publication formats. These findings only characterize published discourse, not AI capability, clinical deployment, or whether the expressed stances were correct.

### Interpretation and Comparison With Prior Work

No prior study has measured the evaluative stance of the medical-AI journal literature directly, so our estimates cannot be benchmarked against earlier work. Bibliometric and scoping studies mapped the field’s growth and topics (9), and the closest stance-adjacent work measured public sentiment toward health AI (11) or the general positivity of scientific abstracts (12, 13); our finding that favourable framing still dominates while unqualified promotion recedes aligns with that work rather than replicating it.

### Temporal Change in Evaluative Stance

The temporal pattern does not support a broad turn against AI. The clearest change was the reduction of unqualified promotion: explicit Advocacy fell from 2.9% in 2021 to 0.6% in 2026 (Spearman ρ=−1.00), while qualified favourable framing remained dominant and explicit scrutiny increased only modestly. This change could reflect composition rather than framing, because the corpus contained more system-testing papers (evaluative) over time. However, critical share rose within both prefilter genres, so composition alone does not explain it (Figure S6, Table S14). The change is real but limited, and mainly one of register: from broad promotion toward more qualified assessment.

The exploratory forward-validation is consistent with this decrease at the level of individual predictions. The boldest early claims fared worst: five of seven Tier-1 superiority-or-replacement claims were not borne out and none were confirmed, and only two of 78 claims were decisively borne out. Because the analysis is Advocacy-only and single-rater, it can show when optimistic predictions over-promised (not borne out), but not whether critical predictions that escaped contradiction were ultimately borne out.

### Changing Content of Critical Discourse

The content of criticism also became more concrete: error and patient-safety concerns occupied more of the critical discourse, while regulation and ethics/bias occupied less. The latter themes did not necessarily decline in volume; their share fell as the critical corpus expanded (Figure S2). A falling share therefore reflects relative emphasis, not the absolute amount of attention in the literature.

The hallucination theme also requires careful consideration. In natural-language-generation research, hallucination can include several forms of unsupported, unfaithful, or incorrect output (6). Our theme therefore included both factual error and fabrication. The mechanism analysis found factual error more often than fabrication, even as generative-model papers became more common. This suggests that published criticism focused more on whether outputs were correct than on whether models invented content. The interpretation remains provisional. Human–model agreement for failure mode was moderate, and the classifier assigned fabrication more readily than the human coder. The estimated fabrication share should therefore be treated as an upper bound.

### Variation Across Specialties, Geography, and Publication Formats

The specialty, geography, and publication-format patterns show that scrutiny was uneven, but they do not explain why. Earlier regulatory studies found that AI/ML-enabled medical-device approvals were concentrated in a small number of specialties, especially radiology (16, 17). We found an inverse association between critical stance and mapped FDA device counts (Spearman ρ=−0.65, two-sided p=0.004). Dermatology was an exception, with a high critical rate despite no mapped cleared devices, which underscores that the FDA count reflects US cleared-device availability, not adoption, deployment, maturity, or clinical use. The direction persisted after specialties with structural zeros were removed, but the reduced analysis was no longer statistically significant at n=11 (Table S5). The analysis was cross-sectional, and therefore cannot establish a mechanism.

Several explanations of this clearance–scrutiny gradient are possible, and we offer them only as conjecture. Familiarity with fielded systems may temper alarm, so that specialties with more cleared devices write less critically. Alternatively, scrutiny may concentrate where systems are least validated for routine use, so that criticism is heaviest where clearance is sparse. A third possibility is that unmeasured specialty factors, such as task type or evidence conventions, shape both criticism and clearance. The cross-sectional design cannot distinguish these accounts, and we do not attempt to.

First-author geography similarly indexes publication metadata and discourse, not national attitudes, research quality, or AI capability. Specialty-mix adjustment explained little of the regional contrast, but publication norms, language, selection, and other unmeasured factors may contribute. Publication-type differences were also descriptive. Reviews were less critical than primary research and opinion formats. One legible reason is theme emphasis: reviews foreground governance and ethics, whereas primary research foregrounds concrete error and safety, and within reviews the more favourable narrative subtype outnumbers the more critical systematic subtype (Tables S9, S10). Outside the medical-AI literature, clinical reviews have attracted more early page views than primary papers, and reviews have been overrepresented among highly downloaded articles in general medical and radiology journals (18–20). These studies were not specific to our corpus and did not identify practising clinicians, but they suggest that publication format may affect the evaluative material readers encounter.

### Implications

The results support more precise language in medical-AI reporting. Favourable conclusions can remain favourable while stating limits, uncertainty, and specific failure modes. Critical reporting should also distinguish factual error from fabrication instead of using hallucination as a single category. Calling both “hallucination” hides whether a model gave a wrong answer, or invented one.

The distribution of scrutiny also matters for research communication. A reader may meet a different evaluative register across specialties and publication formats even when the topic is similar. Which editorial conventions, evidence standards, and article purposes drive that variation is worth closer study. The geographic findings likewise identify variation for qualitative follow-up, not a ranking of countries’ attitude.

### Strengths and Limitations

Every LLM measurement step was validated against blinded human coding, not only the stance classifier. Agreement was substantial for stance (quadratic-weighted κ=0.79) and specialty (κ=0.75), almost perfect for model type (κ=0.84), and moderate for failure mode (κ=0.57) and the prefilter (κ=0.51). The study also separated factual error from fabrication and tested several alternative explanations for the temporal pattern. These features support a reproducible description of published discourse, but they do not remove measurement error. The two weaker steps bound what can be claimed: the prefilter decides which papers enter the corpus, and the failure-mode axis carries the accuracy-over-fabrication reading.

The analysis was limited to titles and abstracts. Abstracts compress findings and may not reflect qualifications in the full text. The classifiers were LLMs, and validation depended mainly on one human coder. The prefilter favoured precision over recall, so the corpus is an incomplete sample of AI-related publications. Temporal filtering concerns were partly addressed by the keep-rate audit (Table S15) and the within-genre analysis (Figure S6, Table S14), but specialty-specific retention could not be assessed because dropped application records carry no specialty label.

The corpus was restricted to English-language, PubMed-indexed Q1/Q2 journals. SJR matching used journal titles and best quartile, which excluded unmatched records and admitted some high-ranking non-medical venues. The 2026 stratum covered January through April and was incomplete. First-author country was inferred from one affiliation and is an approximate proxy for publication geography.

The FDA measure was US-centric and counted cleared devices. Structural zeros reflected the panel mapping, not absence of available AI. Generative-share was calculated among critical papers and was endogenous to criticality. The specialty analysis was cross-sectional. Multiple analyses were descriptive and hypothesis-generating; p values were not adjusted for multiplicity or used for confirmatory decisions. Forward-validation was exploratory, Advocacy-only, single-rater, retrieval-dependent, and limited by the incomplete record through April 2026.

## Conclusion

Medical-AI abstracts shifted away from unqualified promotion and toward more specific evaluation of errors and safety. The change was modest, and Cautious Optimism remained dominant. Published concern focused more on factual accuracy than fabrication, although that result is less certain. Scrutiny varied across specialties, first-author geography, and publication formats.

## Data Availability

All data generated in this study are available online. The analytic dataset, consisting of one row per record with its PubMed identifier and assigned stance, theme, mechanism, specialty, and first-author region labels (n = 16,749), together with all prompts, classification code, mapping tables, and committed model outputs, is available in the public repository at https://github.com/Lixing-Kingsley-Wang/ai-narratives-2026. The underlying bibliographic records were obtained from PubMed and are identified by the PubMed IDs provided in the repository. Raw abstract texts are not redistributed but can be retrieved using the provided code and PubMed IDs.

## Disclaimers

### Use of Artificial Intelligence

Artificial intelligence was used in two roles. First, Claude Haiku 4.5 and Claude Sonnet 4.6 (Anthropic) were used as study instruments for prefiltering, stance classification, thematic classification, mechanism reclassification, and forward-validation claim extraction and neutral summarization, as described in the Methods. Second, Claude Opus 4.6 (Anthropic) assisted with manuscript drafting, editing, and code review.

All model outputs used as data were generated through scripted workflows, checked for schema validity, and used only within the predefined analytic procedures. Human review was performed where specified, including blinded validation and adjudication. AI-generated classifications and summaries were not treated as independent authoritative interpretations beyond their defined analytic roles. No patient-level data were used. AI did not independently determine the study design, statistical analysis plan, substantive interpretation, or final manuscript content - these remained the sole responsibility of the authors.

### Compute and Carbon Footprint

LLM inference was used for prefiltering, stance classification, thematic classification, mechanism reclassification, and forward-validation support. Across Claude Haiku 4.5, Claude Sonnet 4.6, and limited Claude Opus 4.6-assisted workflows, the study used approximately 124.5 million tokens. Under the laboratory compute-carbon calculator’s hosted-inference assumptions, using 0.12 g CO₂e per 1000 tokens, the estimated footprint was approximately 14.9 kg CO₂e. A sensitivity estimate using 0.45 g CO₂e per 1000 tokens yielded 56.0 kg CO₂e {Mozafari et al. 2026, unpublished manuscript}.

Local embedding for retrieval-augmented validation ran for approximately 1 hour on an Apple M2 MacBook Air in Montreal. It contributed a negligible additional operational footprint under the calculator’s Montreal grid-intensity assumption. The LLMs were accessed through hosted services, so provider-side hardware allocation, power usage effectiveness, water use, and data-center location were not directly observable. These values should therefore be treated as workload-level estimates, not direct metered emissions.

### Reflexivity

We began this study expecting the critical stance to peak in 2023, after the first wave of LLM evaluations, and then to settle back toward optimism. The data showed a steady rise instead, and the temporal question changed accordingly: not when criticism peaked, but how far unqualified promotion had receded. One author coded every validation sample and repeatedly adjudicated the boundary between Alarm and Caution, which is where our own reading of hedged conclusions was formed through research rather than applied.

This study also used generative AI as its instrument while analysing how the medical literature evaluates generative AI, so the tool and the object belonged to the same class of system. Two moments made that relation visible. The classifier applied the fabrication label more readily than the human coder, so the very distinction we report, between factual error and fabrication, proved unstable inside our own measurement. And the unvalidated predictive-claim flag manufactured forward-looking predictions from descriptive text in 18 of 92 candidate records, which is why a human gate preceded every verdict. Following Christou (21), we treat reflexivity here as looking forward rather than turning back: analytic agency was distributed between the authors and the models, while accountability for the resulting classifications remains ours. We also acknowledge the constructivist risk he identifies, that delegating classification to a fast and fluent tool can reduce the interpretive friction through which a researcher notices ambiguity. Hand-coding the validation samples was our attempt to keep that friction; we cannot claim it survived intact across 16,749 records.

