## Supplementary Note, Tables and Figures for "Evaluative Stance Toward Artificial Intelligence in High-Quartile Medical Journals (2021– 2026): Large-Scale LLM-Assisted Computational Content Analysis"

### Supplementary Note 1. Data and code availability

All prompts, classification code, mapping tables, PubMed identifiers, and committed analysis outputs supporting this study are available in the public repository:

**https://github.com/Lixing-Kingsley-Wang/ai-narratives-2026**

The analytic dataset, consisting of one row per record with its PubMed identifier and the assigned stance, theme, mechanism, specialty, and first-author region labels (n = 16,749), is provided as Supplementary Data 1 and is also included in the repository. Because hosted large-language-model behaviour drifts over time, the committed model outputs in the repository, rather than re-inference, are the basis for exact reproduction. Analyses were run in Python 3.12; package versions are pinned in requirements.txt.

The table below maps each Methods component to the file(s) that implement it, so any reported number can be located and re-run.

| Methods component | Repository file(s) | Contents |
|---|---|---|
| Corpus retrieval & quartile filtering | fetch_pubmed_medical.py; sjr_filter.py; input/sjr/sjr_2021…2026.csv | Verbatim PubMed query, monthly-window retrieval loop, English/date restrictions, PMID de-duplication, SCImago Q1/Q2 matching |
| Publication-type mapping (5 categories) | robustness.py | PubMed publication-type → 5-category collapse rules |
| Prefilter (isolating AI-engaged literature) | prefilter_batch.py; output/pilot_200.xlsx; output/pilot_200_full_haiku.csv | Verbatim prefilter prompt, KEEP/DROP parsing, and the 200-record human-vs-model pilot |
| Stance classification & validation | classify_stance.py (v1); classify_stance_batch.py (v2); compute_kappa.py; output/validation/*; output/comparison_v1_v2.md | Verbatim stance prompts and few-shot examples, human-coder instructions, full 5×5 confusion matrix, κ at all sensitivity layers, v1→v2 comparison |
| Thematic classification | classify_themes_batch.py | Verbatim ten-theme codebook prompt and multilabel cap logic |
| Mechanism re-classification (failure mode / model type) | classify_q7_failuremode.py; output/analyses/q7_kappa_report_abstractonly.md; q7_validation_* | Verbatim mechanism prompt, calibration examples, both 4×4 confusion matrices, Cohen κ / Gwet AC1 |
| Specialty classification | analysis/classify_specialty.py; output/analyses/specialty_kappa_report.md; specialty_validation_* | 18-category specialty taxonomy prompt, 18×18 confusion matrix, κ |
| First-author geography (8 regions) | analysis/extract_geography.py | Country → 8-region table, alias table, ambiguity-resolution rules |
| FDA cleared-device mapping | output/external/fda_aiml_devices.csv; analysis/run_specialty_aiadoption.py | FDA AI/ML device list (1995–2025) and FDA-panel → specialty mapping, structural-zero list |
| Forward-validation (prophecy panel) | prophecy_phase1_*.py; prophecy_phase2_*.py; output/analyses/prophecy/*; prophecy_panel_preregistration.md | Pre-registered protocol, frozen 74-paper/78-claim manifest, retrieval + adjudication, verdicts with cited evidence PMIDs |
| Statistics & robustness checks | robustness.py (bootstrap_ci); analysis/run_country_critical_map.py; analysis/revision_checks/*.py; output/revision_checks/* | Bootstrap CI method (n = 1000, seed 42) and the sensitivity analyses reported in Tables S5, S8, S14, S15 and Fig. S6 |

### Supplementary Figures

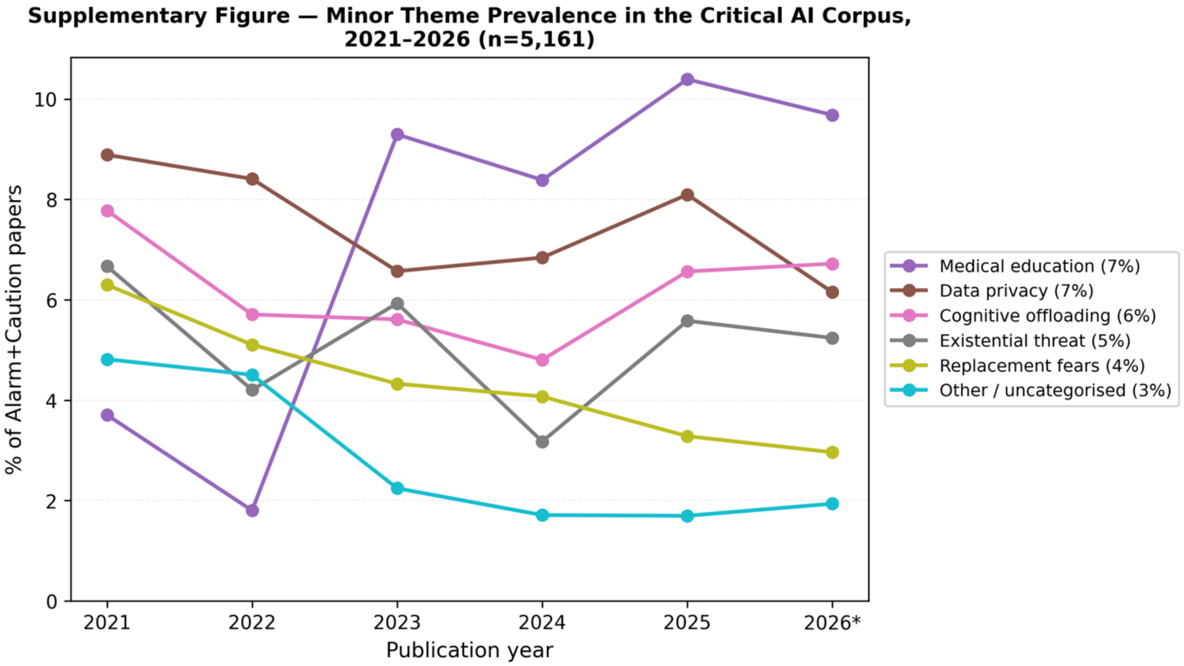


**Figure S1. Prevalence of the six minor concern themes in the critical corpus, 2021–2026.**

*The six themes aggregated as "other" in Figure 1B, shown individually: medical education, data privacy, cognitive offloading, existential threat, replacement fears, and other/uncategorised. Denominator: the 5,161 critical papers; themes are multilabel. Lines are annual point estimates. The percentage beside each label is the mean of the six annual prevalences; pooled prevalences and per-theme Spearman ρ/p are in Table S1. 2026 is partial.*

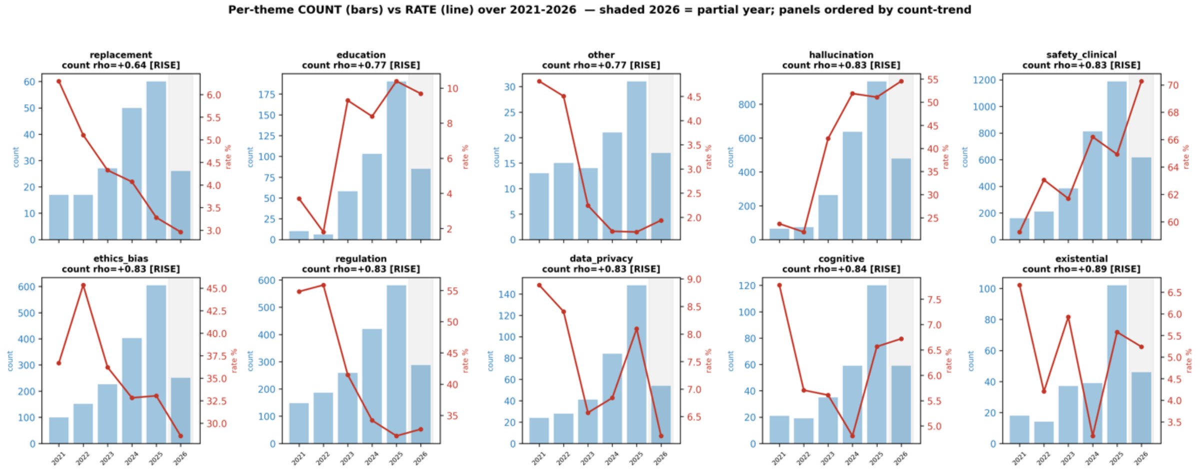


**Figure S2. Absolute count versus prevalence share for all ten themes, 2021–2026.**

*For each theme, the annual paper count (bars) and annual prevalence share (line) within the critical corpus (n=5,161). The governance-related themes (regulation, ethics/bias) declined in prevalence share while their absolute counts rose, showing that their apparent decline reflects dilution by the growing critical corpus rather than a fall in volume. The 2026 stratum is shaded (partial year).*

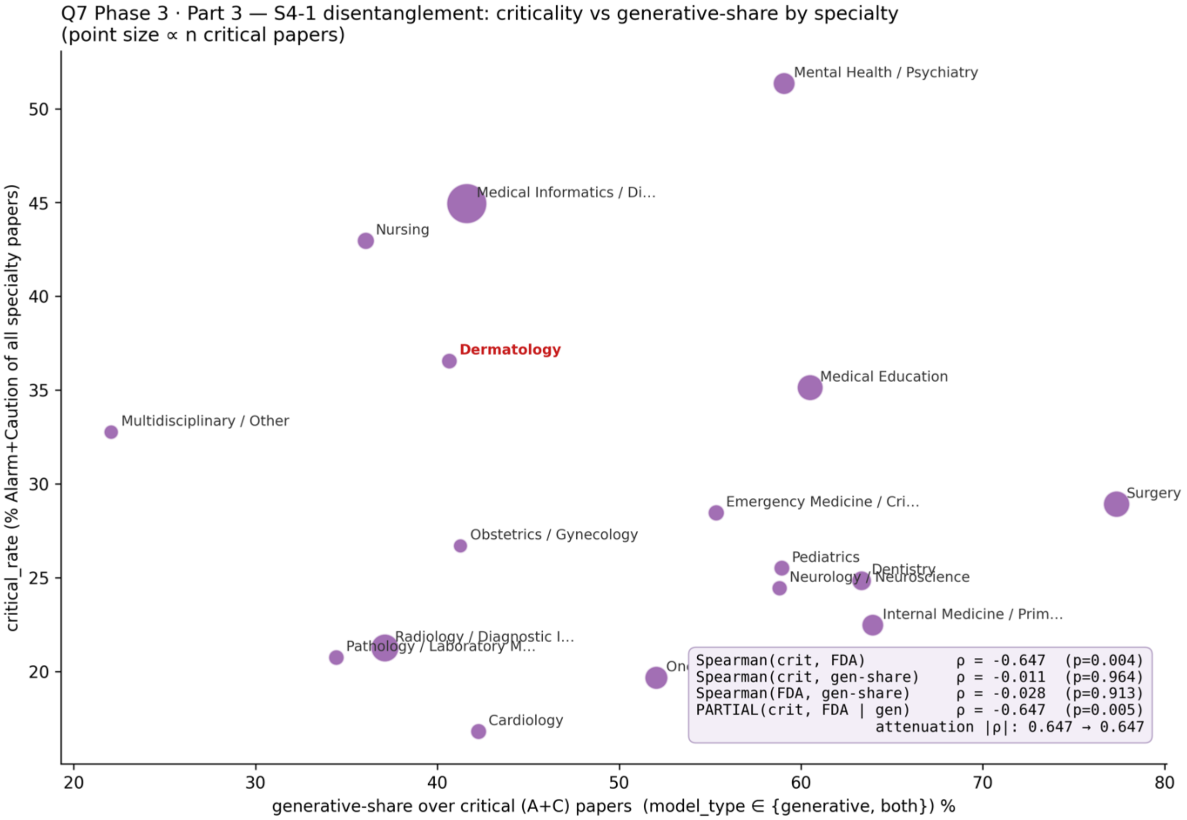


**Figure S3. Critical rate versus generative-model share across specialty/domain categories.**

*Per-category critical rate against generative-share — the proportion of a category's critical papers concerning generative models — with point size proportional to the number of critical papers and Dermatology annotated. Critical rate was essentially uncorrelated with generative-share (Spearman ρ=−0.01) but inversely correlated with FDA cleared-device count (ρ=−0.65, two-sided p=0.004), and the partial correlation controlling for generative-share was unchanged (ρ=−0.65). Generative-share is computed over critical papers only and is therefore endogenous to criticality, so this disentanglement is suggestive. Supports Figure 3B.*

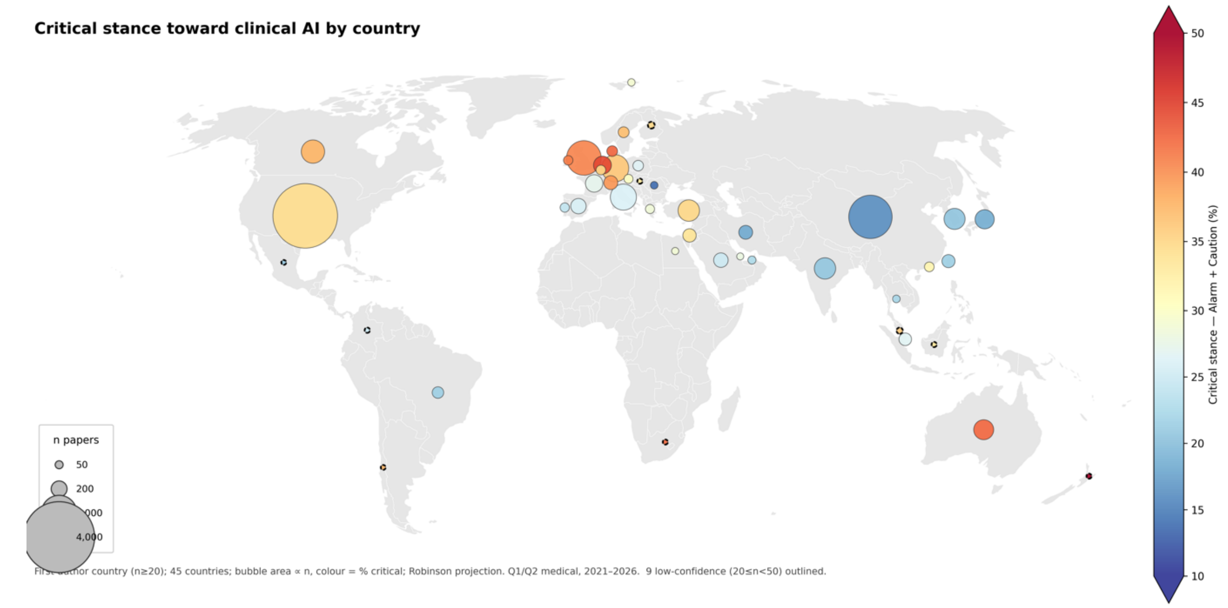


**Figure S4. First-author-country critical-rate map at the ≥20-record threshold.**

*As Figure 4 but at the more permissive cutoff of at least 20 records per country (45 countries); the nine low-confidence countries (20 ≤ n < 50) are outlined. Country is resolved from first-author affiliation only and indexes publication discourse, not national attitudes. Robinson projection; bubble area proportional to the number of papers, fill color the critical rate.*

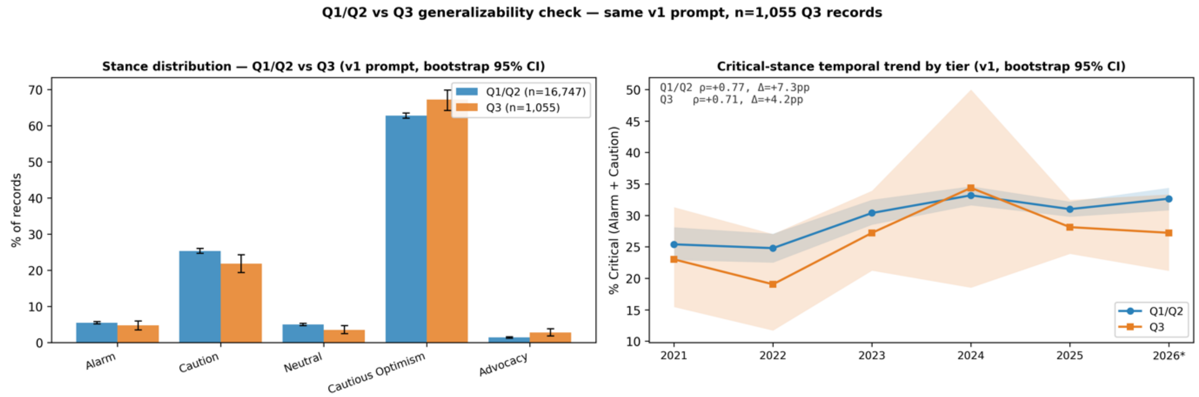


**Figure S5. Quality-tier comparison: Q1/Q2 versus Q3.**

*Left: the five-stance distribution for the Q1/Q2 corpus (n=16,747) versus a Q3 comparison arm (n=1,055) classified with the identical v1 rubric, with 95% percentile bootstrap CIs. Right: the critical-share temporal trend by tier (Q1/Q2 Spearman ρ=+0.77, Δ=+7.3 pp; Q3 ρ=+0.71, Δ=+4.2 pp). Q3 tracks the same temporal direction but is slightly less critical overall; the Q3 2024 stratum is small and reflects an indexing dip. The prefilter-retention audit and within-publication-type standardization are in Table S12. 2026 is partial.*

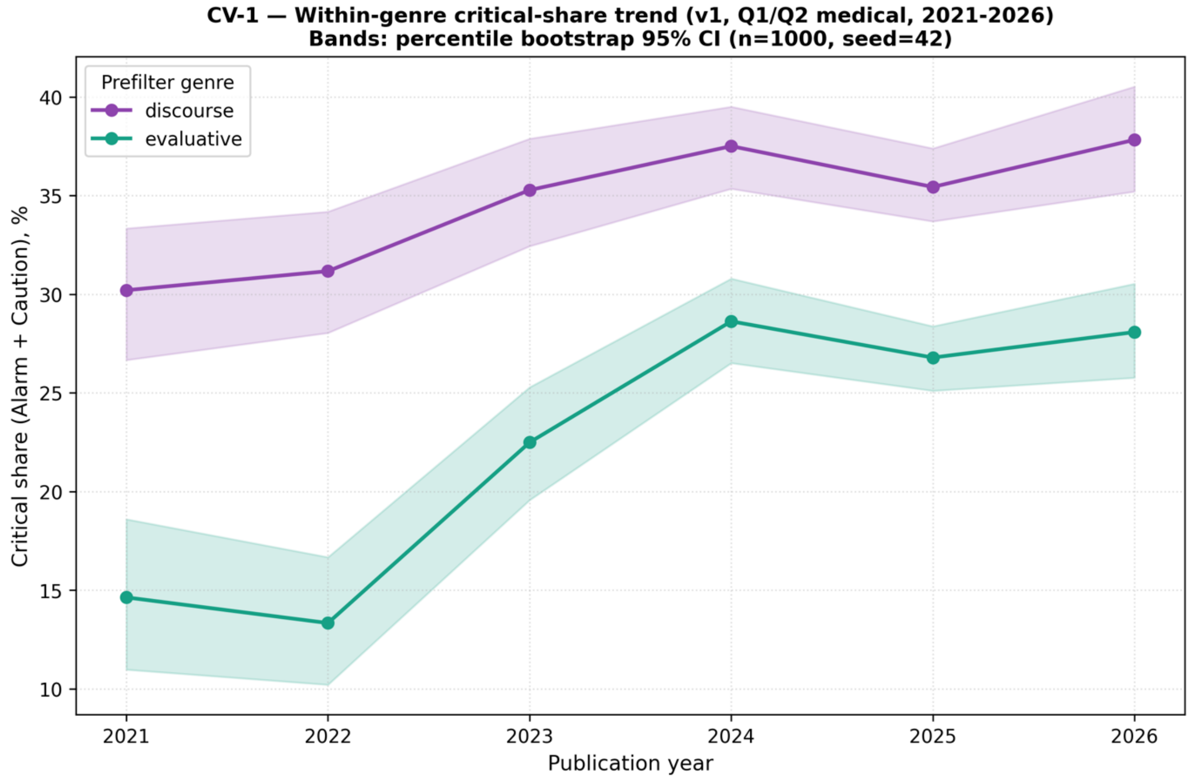


**Figure S6. Within-genre robustness of the critical-share trend.**

*Annual critical share (Alarm + Caution) computed separately within each prefilter genre — discourse and evaluative — with 95% percentile bootstrap CI bands (n=1000, seed=42). The rise reproduces within both genres (discourse 30.2% to 37.8%, Spearman ρ=+0.94; evaluative 14.6% to 28.1%, ρ=+0.77) even though the corpus composition shifted over the period toward the less-critical evaluative genre, so the overall rise is not a genre-composition artifact. Per-year values and the discourse:evaluative ratio are in Table S14.*

### Supplementary Tables

#### Table S1. Per-theme Spearman ρ, all ten themes.

*Share of critical papers (Alarm+Caution, n=5,161) assigned each theme; multilabel, 1–3 per record. ρ = Spearman versus publication year over six annual points. Display term "Patient safety" = code label safety_clinical.*

| Theme | Overall % | 2021 % | 2026 % | Δ pp | ρ |
|---|---|---|---|---|---|
| Patient safety | 65.3 | 59.3 | 70.3 | +11.0 | +0.886 |
| Hallucination / errors | 47.5 | 23.7 | 54.6 | +30.9 | +0.886 |
| Governance / regulation | 36.5 | 54.8 | 32.8 | −22.0 | −0.886 |
| Ethics & bias | 33.6 | 36.7 | 28.6 | −8.1 | −0.886 |
| Medical education | 8.8 | 3.7 | 9.7 | +6.0 | +0.829 |
| Data privacy | 7.3 | 8.9 | 6.2 | −2.7 | −0.771 |
| Cognitive offloading | 6.1 | 7.8 | 6.7 | −1.1 | −0.086 |
| Existential threat | 5.0 | 6.7 | 5.2 | −1.4 | −0.371 |
| Replacement fears | 3.8 | 6.3 | 3.0 | −3.3 | −1.000 |
| Other | 2.1 | 4.8 | 1.9 | −2.9 | −0.829 |

#### Table S2. v1 vs. v2 rubric: per-stance temporal trend.

*v1 canonical; v2 = stricter conclusion-anchored variant. "same/opposite" = trend-direction agreement with v1. Only Cautious Optimism flips, non-significant under both. n=6 permutation floor: p ≥ 0.003 for ρ = ±1.00. Δ = +7.25 pp for Critical uses unrounded annual estimates (rounded endpoints 25.4 → 32.6).*

| Stance | 2021 % | 2026 % | Δ pp | v1 ρ | p | v2 direction (ρ) |
|---|---|---|---|---|---|---|
| Alarm | 2.7 | 6.2 | +3.5 | +0.771 | 0.072 | same (+0.771) |
| Caution | 22.7 | 26.4 | +3.8 | +0.771 | 0.072 | same (+0.714) |
| Neutral | 7.3 | 4.3 | −3.0 | −0.886 | 0.019 | same (−0.943) |
| Cautious Optimism | 64.3 | 62.4 | −1.9 | −0.486 | 0.329 | opposite (+0.486) |
| Advocacy | 2.9 | 0.6 | −2.3 | −1.000 | 0.003 | same (−0.714) |
| **Critical (A+C)** | **25.4** | **32.6** | **+7.25** | **+0.771** | **0.072** | **same (+0.714)** |

#### Table S3. Surviving 2026 Advocacy records (n=17).

*Aggregate mix: 9/17 Medical Informatics, 5/17 explicitly drug-discovery; 11/17 Review; 9/17 North America; predictive-claim 16/17 = 94%.*

| PMID | Journal | Pub type | Specialty | Country | Region | Pred. | Title |
|---|---|---|---|---|---|---|---|
| 41694518 | Frontiers in public health | Review | Multidisciplinary / Other | USA | North America | yes | How AI can be used to promote public and population health. |
| 41276215 | Can. J. of Cardiology | Letter | Pediatrics | Canada | North America | yes | A Custom Artificial Intelligence GPT-4 Model for Pediatric Cardiology: Surpassing the Board Exam Threshold and Transforming Medical Learning. |
| 41499575 | J. of Craniofacial Surgery | Research Article | Surgery | USA | North America | yes | Reimagining Care Delivery in Craniofacial Surgery and Beyond: A Multicenter Analysis of How AI Automation Can Reshape Cost Efficiency in US Health Care Across Specialties. |
| 42039741 | ACS central science | Review | Medical Informatics / Digital Health | USA | North America | yes | From Prompt to Drug: Toward Pharmaceutical Superintelligence. |
| 41206190 | Hand clinics | Review | Medical Informatics / Digital Health | Taiwan | East Asia | yes | Artificial Intelligence-Driven Personalized Medicine. |
| 41392515 | JEADV | Review | Dermatology | Germany | Western Europe | yes | Precision dermatology 2050: AI-driven personalized monitoring and individualized treatment. |
| 41722216 | Acta psychologica | Research Article | Medical Informatics / Digital Health | China | East Asia | no | How personal anticipations and perceived gratifications influence continuous use intention toward AI-driven chatbots? Moderating roles of perceived innovativeness and active involvement. |
| 40865514 | Cell systems | Review | Medical Informatics / Digital Health | USA | North America | yes | Illuminating the universe of enzyme catalysis in the era of artificial intelligence. |
| 41636234 | Chemical Society reviews | Review | Medical Informatics / Digital Health | USA | North America | yes | Artificial intelligence-powered nanomedicine. |

| PMID | Journal | Pub type | Specialty | Country | Region | Pred. | Title |
|---|---|---|---|---|---|---|---|
| 41551828 | World J. of Gastroenterology | Review | Internal Medicine / Primary Care | Colombia | Latin America / Oceania | yes | Artificial intelligence in metabolic dysfunction-associated steatotic liver disease: Transforming diagnosis and therapeutic approaches. |
| 41431407 | Yonsei medical journal | Research Article | Medical Informatics / Digital Health | South Korea | East Asia | yes | How Does Medical Artificial Intelligence Revolutionize Physician Productivity? |
| 41389441 | Pharmacological reviews | Review | Medical Informatics / Digital Health | Australia | Latin America / Oceania | yes | Leading artificial intelligence-driven drug discovery platforms: 2025 landscape and global outlook. |
| 41825996 | Dental clinics of N. America | Review | Dentistry | USA | North America | yes | ProSocial Artificial Intelligence in Oral Health: A Paradigm Shift. |
| 41667642 | Scientific reports | Research Article | Medical Informatics / Digital Health | China | East Asia | yes | Multimodal large language models challenge NEJM image challenge. |
| 41460502 | J. Continuing Educ. Nursing | Research Article | Nursing | USA | North America | yes | AI for Nurses: Did You Know There Is an AI for That? |
| 41826009 | Dental clinics of N. America | Review | Dentistry | USA | North America | yes | The Convergence of Prosthodontics and ProSocial Artificial Intelligence: Advancing Equity and Excellence in Dental Care. |
| 41431184 | Drug development research | Review | Medical Informatics / Digital Health | India | South/Southeast Asia | yes | Mathematical and Artificial Intelligence Techniques in Modern Drug Discovery: A Review. |

#### Table S4. Fabrication share among generative-model papers, by year (A+C).

*Denominator = generative-model critical papers each year (abstract-present). Flat ~20–23% = the Figure 2B plateau. 2021–22 suppressed (generative n too small).*

| Year | Fabrication share among generative papers (%) |
|---|---|
| 2023 | 21.8 |
| 2024 | 22.9 |
| 2025 | 19.7 |
| 2026 | 22.0 |

#### Table S5. FDA clearance–scrutiny gradient: robustness.

*18 specialty/domain categories. Generative-share computed on A+C papers (endogenous to criticality). ρ shown to 2 dp.*

| Relationship | ρ | p (2-sided) | n |
|---|---|---|---|
| Critical rate vs. FDA cleared-device count (zero-order) | −0.65 | 0.004 (1-sided 0.002; perm 0.002) | 18 |
| Critical rate vs. generative-share | −0.01 | 0.964 | 18 |
| FDA count vs. generative-share | −0.03 | 0.913 | 18 |
| Partial: critical rate vs. FDA \| generative-share | −0.65 | 0.005 | 18 |
| critical rate vs. FDA — structural zeros dropped | −0.58 | 0.062 | 11 |
| (sensitivity) Critical rate vs. in-corpus evaluation-share proxy | −0.47 | 0.026 (perm 0.028) | 18 |

#### Table S6. East-Asia per-country critical rate.

*First-author country within East Asia. Anglophone-publishing gradient. 95% bootstrap CI.*

| Country | n | Critical % (95% CI) |
|---|---|---|
| Hong Kong | 95 | 31.6 (21.1–41.0) |
| Singapore | 179 | 26.8 (20.7–33.5) |
| Taiwan | 169 | 21.3 (15.4–27.2) |
| South Korea | 487 | 20.3 (16.6–24.2) |
| Japan | 424 | 19.6 (15.8–23.6) |
| China | 1,904 | 16.2 (14.6–17.8) |

#### Table S7. East-Asia within-specialty Δ (EA − NA/WEU).

| Specialty | n EA | n NA+WEU | EA % | NA+WEU % | Δ pp |
|---|---|---|---|---|---|
| Medical Education | 188 | 507 | 18.1 | 41.0 | −22.9 |
| Medical Informatics | 746 | 2,561 | 31.4 | 50.2 | −18.8 |
| Surgery | 173 | 918 | 17.9 | 31.7 | −13.8 |
| Radiology | 568 | 1,188 | 12.2 | 24.7 | −12.5 |

| Specialty | n EA | n NA+WEU | EA % | NA+WEU % | Δ pp |
|---|---|---|---|---|---|
| Ophthalmology | 200 | 353 | 11.0 | 22.9 | −11.9 |
| Oncology | 368 | 743 | 11.7 | 23.1 | −11.5 |
| Dentistry | 131 | 209 | 16.0 | 27.3 | −11.2 |
| Internal Medicine | 264 | 579 | 14.0 | 25.0 | −11.0 |
| Pathology | 91 | 299 | 13.2 | 23.4 | −10.2 |
| Cardiology | 112 | 397 | 15.2 | 18.1 | −3.0 |

**Table S8. Publication type × stance (full 5×5).**

*Row % (1 dp) across the five stances; critical = Alarm+Caution; favorable = Cautious Optimism+Advocacy.*

| Pub type | n | Alarm | Caution | Neutral | Caut. Opt. | Advocacy | Critical | Favorable |
|---|---|---|---|---|---|---|---|---|
| Research Article | 10,315 | 7.8 | 25.2 | 5.6 | 60.7 | 0.8 | 32.9 | 61.5 |
| Editorial | 404 | 3.2 | 35.2 | 9.4 | 46.0 | 6.2 | 38.4 | 52.2 |
| Review | 5,444 | 1.1 | 23.8 | 2.0 | 71.3 | 1.8 | 24.9 | 73.1 |
| Commentary | 104 | 8.7 | 45.2 | 6.7 | 37.5 | 1.9 | 53.9 | 39.4 |
| Letter | 480 | 7.5 | 34.6 | 21.7 | 30.2 | 6.0 | 42.1 | 36.3 |

**Table S9. Review subtype: systematic vs. narrative.**

*Neutral = remainder to 100%. Narrative subtype carries the format's positivity (7× the Advocacy rate).*

| Subtype | n | Critical % | Alarm % | Caution % | Caut. Opt. % | Advocacy % |
|---|---|---|---|---|---|---|
| Systematic Review | 980 | 29.3 | 3.0 | 26.3 | 68.2 | 0.3 |
| Review (narrative) | 4,464 | 23.9 | 0.7 | 23.2 | 72.0 | 2.1 |

**Table S10. Theme emphasis by format (within A+C).**

| Theme | Reviews % | Research % | Δ pp |
|---|---|---|---|
| Regulation | 57.4 | 27.0 | +30.5 |
| Ethics & bias | 44.0 | 29.4 | +14.6 |
| Data privacy | 10.9 | 6.1 | +4.9 |
| Patient safety | 65.8 | 64.7 | +1.0 |
| Other | 3.3 | 1.7 | +1.5 |
| Replacement | 2.7 | 3.9 | −1.2 |
| Existential | 3.9 | 5.1 | −1.2 |
| Cognitive | 4.4 | 6.6 | −2.1 |
| Education | 5.5 | 10.3 | −4.7 |
| Hallucination | 30.2 | 54.3 | −24.2 |

**Table S11. Forward-validation: Tier × verdict matrix.**

*N = 78 claims (74 papers). Margins verified: not-borne 8; too-early 14. Corpus-silent (9: T1=0/T2=8/T3=1) folds into too-early.*

| Tier (n) | borne_out | partially | not_borne_out | too_early / unfalsifiable |
|---|---|---|---|---|
| Tier 1 — superiority/replacement (7) | 0 | 1 | 5 | 1 |
| Tier 2 — capability deployment (36) | 2 | 23 | 1 | 10 |
| Tier 3 — diffuse transformation (35) | 0 | 30 | 2 | 3 |
| **Total** | **2** | **54** | **8** | **14** |

**Table S12. Q3 vs. Q1/Q2 stance distribution (quality-tier arm).**

*Q1/Q2 n=16,749; Q3 n=1,055, same v1 prompt. Caveat: Q3 2024 n=32 (indexing dip). Percentages 1 dp.*

| Stance | Q1/Q2 | Q3 | Δ pp |
|---|---|---|---|
| Alarm | 5.5 | 4.7 | −0.7 |
| Caution | 25.4 | 21.8 | −3.6 |
| Neutral | 5.0 | 3.5 | −1.5 |
| Cautious Optimism | 62.8 | 67.2 | +4.4 |

| Stance | Q1/Q2 | Q3 | Δ pp |
|---|---|---|---|
| Advocacy | 1.4 | 2.8 | +1.4 |
| **Critical (A+C)** | **30.8** | **26.5** | **−4.3** |

#### Table S13. Industry vs. academic first-author stance.

*First-author affiliation heuristic (~15–20% false positives on generic corporate suffixes). Directionally ~3 pp less critical; CIs overlap. Pub-type-adjusted industry = 27.3%.*

| Group | n | Critical % (95% CI) |
|---|---|---|
| Industry | 285 (1.7%) | 27.7 (22.8–33.0) |
| Academic | — | 30.8 (30.0–31.5) |
| Any-industry (incl. mixed) | 1,209 | 27.7 (25.1–30.4) |

#### Table S14. Within-genre critical share + genre ratio.

*Critical share (Alarm+Caution) by prefilter genre, v1, 2021–2026, with 95% bootstrap CIs. The rise reproduces within both genres — a Simpson's-paradox check that passes.*

| Year | Discourse crit % (95% CI) | Discourse n | Evaluative crit % (95% CI) | Evaluative n | disc:eval |
|---|---|---|---|---|---|
| 2021 | 30.2 (26.7–33.3) | 735 | 14.6 (11.0–18.6) | 328 | 2.24 |
| 2022 | 31.2 (28.0–34.2) | 863 | 13.3 (10.2–16.7) | 480 | 1.80 |
| 2023 | 35.3 (32.4–37.9) | 1,267 | 22.5 (19.6–25.3) | 787 | 1.61 |
| 2024 | 37.5 (35.3–39.5) | 1,901 | 28.6 (26.5–30.8) | 1,799 | 1.06 |
| 2025 | 35.4 (33.7–37.4) | 2,867 | 26.8 (25.1–28.4) | 3,031 | 0.95 |
| 2026 | 37.8 (35.2–40.5) | 1,261 | 28.1 (25.8–30.5) | 1,428 | 0.88 |

#### Table S15. Prefilter keep-rate audit by year.

*KEEP = discourse+evaluative retained by the prefilter; denominator = pre-prefilter eligible Q1/Q2 records dated 2021–2026. v1. Rates 1 dp.*

| Year | kept | eligible | KEEP % |
|---|---|---|---|
| 2021 | 1,064 | 11,191 | 9.5 |
| 2022 | 1,344 | 13,136 | 10.2 |
| 2023 | 2,055 | 14,128 | 14.6 |
| 2024 | 3,704 | 19,372 | 19.1 |
| 2025 | 5,899 | 28,873 | 20.4 |
| 2026 | 2,691 | 10,757 | 25.0 |
| **All** | **16,757** | **97,457** | **17.2** |

#### Table S16. Stance-classifier run-to-run stability, T=1.0.

*Same 300 validation records classified twice at production default T=1.0; canonical v1 labels and human codes untouched. [Editorial note: confirm keep/drop with Dan before submission.]*

| Metric | Value |
|---|---|
| Records (both runs valid) | 300 / 300 (0 FAILED) |
| Raw agreement | 95.3% (286/300) |
| Quadratic-weighted Cohen κ | 0.974 |
| Labels shifted | 14 — all adjacent, no polar flips |
